# Vital signs, demographics, and clinical events for low-birth-weight infants from four intensive care units

**DOI:** 10.64898/2026.04.15.26350178

**Authors:** Ian German Mesner, Doug E. Lake, Sherry L. Kausch, Katy N. Krahn, Angela Gummadi, Timothy W. Clark, Justin C. Niestroy, Rakesh Sahni, Zachary A. Vesoulis, David B. Gootenberg, N. Ambalavanan, Colm P. Travers, Karen D. Fairchild, Brynne A. Sullivan

## Abstract

Premature very low birth weight (VLBW) infants have high rates of mortality and morbidity from sepsis, necrotizing enterocolitis, and respiratory failure requiring intubation and mechanical ventilation. Earlier detection of cardiorespiratory deterioration using vital signs from continuous physiological monitoring may lead to more timely interventions and improved outcomes. To further this research area, we present PreMo, a publicly available dataset of continuous heart rate and oxygen saturation, demographics, clinical events, and outcomes for 3,829 VLBW patients from four Neonatal Intensive Care Units (NICUs) in the United States. The PreMo dataset consists of a collection of parquet files, RO-Crate metadata, and sample usage code scripts hosted on the University of Virginia LibraData Dataverse website.

## Background & Summary

Premature birth remains a critical public health challenge, with preterm infants facing substantially elevated mortality and morbidity rates compared to full-term newborns.^1^ Among the leading causes of death in preterm infants during hospitalization in the NICU is sepsis, a life-threatening systemic response to infection that poses particular risks in this vulnerable population with immature organ systems. Another important disease in preterm NICU patients is necrotizing enterocolitis (NEC). Fortunately, the incidence of both sepsis and NEC have declined with increased breast milk feeding and improved hand hygiene and central line practices, but these illnesses continue to cause substantial morbidity and mortality. The early detection and management of sepsis and NEC in premature infants is complicated by the often subtle and non-specific nature of initial clinical presentations.^2^ This was the impetus for the development of predictive analytics from continuous physiological monitoring to identify at-risk patients.^3,4^ Display of a heart rate characteristics score reflecting imminent sepsis risk was shown to reduce all-cause mortality and sepsis-associated mortality among VLBW preterm infants^.5,6^

Vital sign monitoring has emerged as a valuable tool for sepsis surveillance in the ICU setting, including the NICU. Characteristic patterns in heart rate, respiratory rate, oxygen saturation, and other physiological parameters have been shown to precede clinical sepsis diagnoses, providing opportunities for earlier intervention.^7^ Building on these observations, research has demonstrated that heart rate characteristics monitoring and derived risk scores can successfully reduce mortality when integrated into clinical decision-making.^8^ More recently, advanced computational approaches have extended beyond traditional heart rate analysis to incorporate multi-modal vital sign features for prediction of not only sepsis but also respiratory complications and overall mortality risk in preterm infants.^9,10^ Specifically, our group has shown that adding analysis of systemic oxygenation saturation from pulse oximetry (SpO2) improves on HR-only models for predicting imminent, actionable clinical deterioration.

Despite promising developments in predictive analytics for adverse neonatal events and outcomes, progress in the field has been hindered by limited access to large-scale, multi-institutional datasets that capture the complexity and diversity of NICU patient populations and the continuous-monitoring infrastructure that support them. The PreMo (**Pre**dictive **Mo**nitoring in Premature Infants) multisite dataset aims to address this gap by providing high-resolution (1hz–0.2hz) heart rate (HR) and pulse oximetry (SpO_2_) vital signs along with demographic information, clinical events, and associated outcomes of 3,829 patients from four level IV NICUs (Table 1).

**Table 1:**
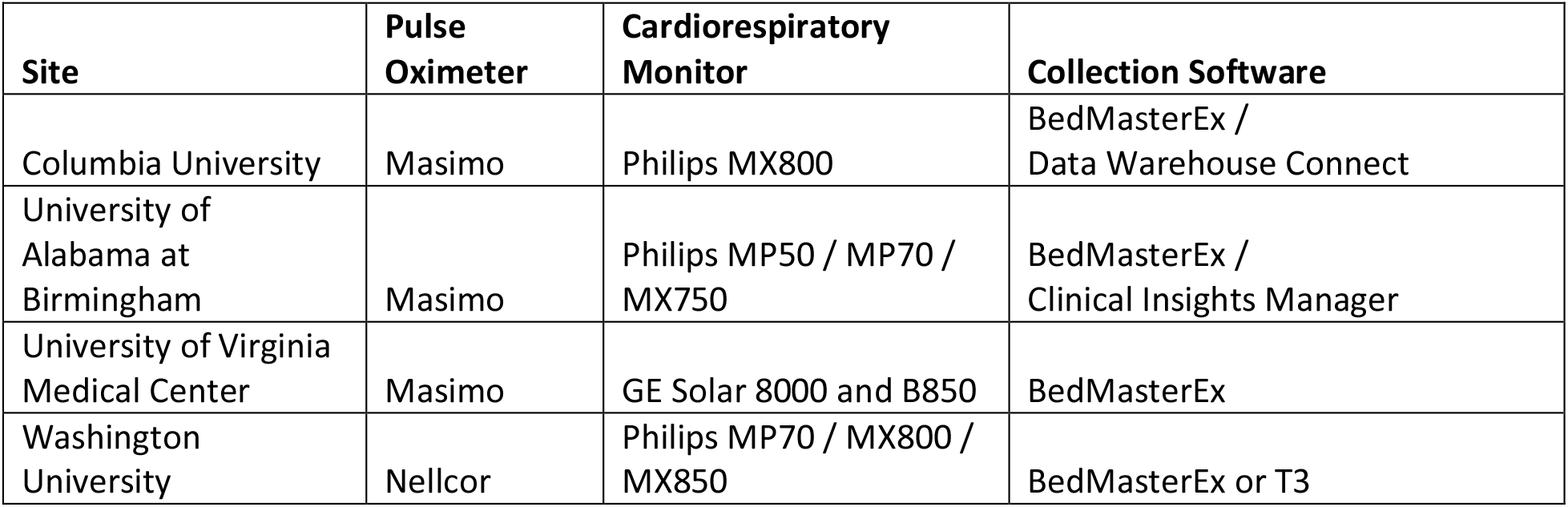
Standard bedside monitors and vital sign data collection software by site.

| Site | Pulse Oximeter | Cardiorespiratory Monitor | Collection Software |
| --- | --- | --- | --- |
| Columbia University | Masimo | Philips MX800 | BedMasterEx /<br>Data Warehouse Connect |
| University of Alabama at Birmingham | Masimo | Philips MP50 / MP70 / MX750 | BedMasterEx /<br>Clinical Insights Manager |
| University of Virginia Medical Center | Masimo | GE Solar 8000 and B850 | BedMasterEx |
| Washington University | Nellcor | Philips MP70 / MX800 / MX850 | BedMasterEx or T3 |

This dataset has been used internally as the basis for a substantial body of NICU research by members of an NIH-funded collaboration (R01HD072071), including risk modeling of late-onset sepsis and NEC^9,11^, identifying clinical events associated with sepsis and NEC^12^, quantifying the impact of bronchopulmonary dysplasia on tachycardia-desaturation events^13^, discovering vital sign patterns associated with dexmedetomidine administration^14^, and others. Furthermore, the detailed clinical and demographic data included have been used to test associations between socioeconomic indicators and adverse outcomes.^15^

Publication of this deidentified dataset advances the FAIR (Findable, Accessible, Interoperable, and Reusable) data principles^16^ within neonatal critical care research, enabling independent validation, replication, and extension of existing models, analytical methods, and scientific findings. It also provides extensive AI-readiness metadata^17^ for this dataset in FAIRSCAPE RO-Crate packages^18,19^.

Although the dataset is large and uniquely integrates clinical and physiological data, several limitations should be acknowledged. Data collection was primarily focused on baseline characteristics and blood culture–related events, reflecting the original NIH-funded emphasis on late-onset neonatal sepsis and NEC. Data collected for secondary focuses are incomplete, including for intubation events, 2-year neurodevelopmental outcomes, and the Area Deprivation Index^20^. The database evolved across successive projects, with data fields added or removed over time, and automated EHR data extraction was not uniformly available across participating sites, limiting the overall scope of available variables. Despite these constraints, all included data elements were carefully curated, annotated, and clinically reviewed, preserving data integrity while maintaining meaningful clinical context. By making this rigorously curated resource openly available, this dataset lays a foundation for collaborative discovery and innovation, accelerating progress toward more precise, data-driven approaches to improving outcomes for critically ill newborns.

## Methods

The PreMo project is an NIH funded (grant R01HD072071) collaboration between four NICUs: University of Virginia (UVA) Health Children’s, Columbia University’s (CU) New York-Presbyterian Morgan Stanley Children’s Hospital, St. Louis Children’s Hospital/ Washington University (WU), and University of Alabama at Birmingham (UAB) (Table 1). Patient demographics, clinical events, and outcomes are manually curated by clinicians and research coordinators in a REDCap database^21,22^ administered at UVA (Table 2). HR and SpO2 data were collected from standard NICU bedside monitors (GE or Philips). Pulse oximetry technology was Masimo (Masimo, Irvine, CA) at 3 sites and Nellcor (Medtronic, Boulder, CO) at 1 site, with default SpO2 averaging (typically 8 seconds). Data were centrally processed and stored using BedMasterEx (Hilrom, Chicago, IL), T3 (Etiometry, Boston, MA), Data Warehouse Connect (Philips Medical, Andover, MA), or Clinical Insights Manager (Philips Medical, Andover, MA). Patients were mapped to beds using a variety of automated and manual processes, including admit discharge transfer (ADT) tracking and device association.

**Table 2:** REDCap form descriptions. See metadata.parquet for field descriptions.

| REDCap form | Rows | Cols | Description |
| --- | --- | --- | --- |
| Demographic and Perinatal | 6,837 | 34 | -Patient demographics: gestational age, birthweight, race, ethnicity, sex<br>-Social determinants of health: Area Deprivation Index<br>-Perinatal variables: Apgar scores, delivery mode, antenatal steroids, admission type (inborn or outborn) |
| Blood Culture Events | 8,139 | 116 | Sepsis evaluations at >3 days of age: ages at blood or other fluid cultures, results, organism, diagnosis, duration of antibiotics administration |
| Intubations | 2,552 | 14 | Age and indication for intubation, age at extubation, |
| Discharge & Outcomes | 6,553 | 28 | - Intraventricular hemorrhage grade<br>-Treated retinopathy of prematurity<br>-Supplemental oxygen at 36 weeks postmenstrual age<br>-Death before NICU discharge<br>-Age at NICU discharge or hospital transfer,<br>-Bayley Scale of Infant Development exam at 2 years of age |

### Study Population and Data Collection

Data were collected on infants admitted to participating NICUs that were VLBW, defined as birth weight less than 1500 grams. These meet the World Health Organization International Classification of Diseases criteria for very low birth weight (ICD-10 P07.00-03 and P07.14-15). Infants were classified as *inborn* if delivered at the admitting hospital’s labor and delivery unit, or *outborn* if transferred after birth from another facility.

### Demographic and Perinatal Characteristics

Maternal race and ethnicity were recorded as self-identified in the medical record or clinical databases. Neighborhood socioeconomic status was assessed using the National Area Deprivation Index (ADI) percentile^20^ derived from the address associated with the NICU admission encounter and 2023 ADI national percentiles.^23^ Gestational age at birth, birth weight, mode of delivery, and multiple gestation status were recorded. Perinatal characteristics included Apgar scores at 1, 5, and 10 minutes (when assigned), antenatal steroid administration (defined as at least one dose of betamethasone given prior to delivery), and membrane rupture duration. Indication for delivery was categorized into preterm labor (including preterm premature rupture of membranes), uteroplacental insufficiency (including intrauterine growth restriction or abnormal umbilical artery Doppler findings), fetal indication (including fetal heart rate concerns or low biophysical profile), maternal indication (including pre-eclampsia), and other indications as documented in the admission note. Clinical chorioamnionitis was recorded when noted in the neonate’s admission documentation.

### Events: Blood Cultures and Intubations

We systematically captured blood culture events at >72 hours from birth for evaluation of suspected late-onset sepsis or NEC. Each unique blood culture event was defined as a blood culture obtained for clinical signs of sepsis or NEC and at least three calendar days since antibiotics were discontinued if there was a recent prior event. The time of each event corresponded to when the blood culture was drawn. For infants with multiple blood culture events throughout the NICU stay, all positive blood culture events were recorded, along with negative blood culture events treated with at least 5 days of antibiotics with final diagnosis of NEC, urinary tract infection, pneumonia, or clinical sepsis. Additionally, blood culture events treated with fewer than 5 days of antibiotics were recorded as sepsis ruled out if negative, contaminant if positive, or a diagnosis for which treatment was cut short due to death.

For each blood culture event, we recorded whether positive, organism, days of intravenous (IV) antibiotic treatment, and associated cultures obtained during the same evaluation. When available, viral testing results around the time of the blood culture were also collected. Treatment duration was recorded as a continuous variable (days of IV antibiotics) and as a binary variable (yes/no treated for five or more consecutive days with any IV antibiotic used for empiric or targeted treatment). For a subset of events included in a published study^2^, more detailed clinical data were recorded at the time of the sepsis evaluation. The variables recorded for this subset of blood culture events included respiratory support device, vascular access, feeding status, feeding type, and clinical signs prompting the sepsis evaluation. Respiratory support options included conventional ventilation, high-frequency oscillatory ventilation, high-frequency jet ventilation, neurally adjusted ventilatory assist, non-invasive positive pressure ventilation, continuous positive airway pressure, high flow nasal cannula, and low flow nasal cannula. Feeding status was recorded as NPO, advancing, or full volume, and feed type was recorded as breast milk, formula, or both.

Intubations were recorded if they occurred between 3 and 120 days, were followed by at least 24 hours of mechanical ventilation, and were preceded by at least 72 hours without mechanical ventilation. The indication for intubation was recorded as apnea, respiratory distress, surfactant administration, surgery/procedure, or other. Intubations for surfactant administration alone or for a surgery or procedure were excluded. The age at intubation and extubation was recorded for each event.

### Neonatal Outcomes

Outcomes diagnosed before NICU discharge were recorded as follows: highest grade of intraventricular hemorrhage (IVH) documented on cranial ultrasound, based on the radiologist’s read using Papile staging criteria^24^; supplemental oxygen requirement at 36 weeks postmenstrual age (inspired oxygen fraction greater than 21%); and severe retinopathy of prematurity (ROP), defined as ROP requiring treatment with laser photocoagulation or anti-vascular endothelial growth factor therapy. Discharge data included length of initial hospitalization and disposition (home, transfer to another facility, or death), Cause of death was documented when known. Neurodevelopmental follow-up data at 2 years of age from Bayley Scales of Infant Development examinations (BSID III or IV)^25,26^ were collected when available and recorded as the composite score across the cognitive, motor, and language components of the exam.

### Deidentification

Patient deidentification in accordance with the HIPAA safe harbor standard^27^ involved removal of all existing patient identifiers (including medical record numbers and other clinical identifiers) and dates associated with birth, clinical events and vital signs. All datasets are instead linked through a new *subject_number_deid* patient identifier, and all dates have been converted to ages. Additionally, the following were removed to help protect from reidentification: most free text fields, multiple-birth linkages and race attributes for small groups (15 patients of *American Indian or Alaska Native* and *Native Hawaiian or Other Pacific Islander*). Birth year was preserved for trend analysis, but was likewise omitted for small groups (5 patients at two sites between 2012-2015). Birth time was converted to UTC and truncated to 10-minute intervals. Site identifiers were anonymized and are referred to as sites 1-4. All other forms of protected health information such as postal addresses and social security numbers were already absent from dataset sources. The resulting deidentified data were internally reviewed for strict adherence to all federal law and organizational procedures and deemed acceptable for public release.

### Ethics statement

Institutional Review Boards at each center approved data collection for retrospective analysis with waiver of consent. Protocol identifiers by site follow:

- Columbia University: IRB Exp, #IRB-AAAP5807
- University of Alabama at Birmingham: #IRB-300007524
- University of Virginia: IRB-HSR #21237
- The Washington University in St. Louis: #201803222

### Data Records

Data are made available publicly as downloadable parquet files on the University of Virginia’s LibraData website^28^, itself a Dataverse instance.^29^ Each REDCap form (Table 2) is exported into a parquet file (e.g. *‘redcap*.*Demographic and Perinatal data*.*parquet’*), and all vital signs (HR and SpO_2_) are exported into a Delta Lake parquet dataset^30^ with one file per patient, each having the same schema (Table 3). REDCap form fields are described in *redcap*.*metadata*.*parquet*, with the exception of columns created as part of the deidentification process including *subject_number_deid* and all columns with a suffix of *_age_us* where dates were converted to ages. These age columns were written as parquet *duration* types for REDCap files and 64-bit integer timestamps for vitals files. Additionally, RO-Crate metadata is included in *ro-crate-metadata*.*json*.

**Table 3:** Schema of patient HR and SpO_2_ data in parquet files.

| Column | Type | Description |
| --- | --- | --- |
| site_id | Integer | Site identifier (1-4, anonymized) |
| subject_number_deid | Integer | Patient identifier (7 digits, deidentified) |
| age_us | Integer | Age in microseconds |
| HR | Float | Electrocardiogram-derived heart rate in beats per minute (0-300). Frequency and technical definition can vary by site, device, and processing software. |
| SPO2 | Float | Plethysmography-derived oxygen saturation (0-100). Frequency and technical definition can vary by site, device, and processing software. |

### Data Overview

Demographics and outcomes from REDCap are present for 6,837 patients, and 3,829 of these have over 10 billion HR and/or SpO2 records spanning over 4 million patient-hours. A summary of demographics, outcomes, and vital sign availability by site is presented as a *gtsummary* table^31^ in Table 4, and a sample hour length segment of HR and SpO_2_ is plotted in Figure 1. Vital sign data were not available for all infants due to technical limitations, so the number of infants with any vital sign data is lower than the overall cohort.

**Table 4:** gtsummary^31^ of demographics, events, outcomes, and vital sign availability for PreMo patients. All counts represent number of patients. ‘Birth Weight’ is reported in grams, and ‘Gestational Age’ in weeks. See text for intubation inclusion criteria. ‘Any Vital Signs’ indicates existence of a parquet file with HR and SpO_2_. ‘Vital Signs (hours of life)’ reports the number of patient hours of life with any HR or SpO_2_ signal, while ‘Vital Signs(coverage)’ is this metric relative to the number of hours between birth and discharge. A script to reproduce this table—with the exception of vital sign availability metrics—is included in the dataset submission^28^.

| **Characteristic** | **Site 1**<br>N = 1,623<sup>†</sup> | **Site 2**<br>N = 1,484<sup>†</sup> | **Site 3**<br>N = 2,160<sup>†</sup> | **Site 4**<br>N = 1,570<sup>†</sup> |
| --- | --- | --- | --- | --- |
| Female | 795 (49%) | 716 (48%) | 1,084 (50%) | 808 (51%) |
| Race |  |  |  |  |
| Asian | 16 (1.0%) | 22 (1.5%) | 131 (6.1%) | 20 (1.3%) |
| Black | 385 (24%) | 552 (37%) | 745 (34%) | 772 (49%) |
| Unk/Other | 163 (10%) | 46 (3.1%) | 371 (17%) | 81 (5.2%) |
| White | 1,059 (65%) | 864 (58%) | 913 (42%) | 697 (44%) |
| Ethnicity |  |  |  |  |
| Latino | 135 (8.3%) | 59 (4.0%) | 469 (22%) | 119 (7.6%) |
| Non-Latino | 1,336 (82%) | 1,387 (93%) | 709 (33%) | 1,421 (91%) |
| Unk/Other | 152 (9.4%) | 38 (2.6%) | 982 (45%) | 30 (1.9%) |
| Birth Year |  |  |  |  |
| 2012-2016 | 541 (33%) | 199 (13%) | 743 (34%) | 0 (0%) |
| 2017-2021 | 580 (36%) | 808 (55%) | 919 (43%) | 441 (28%) |
| 2022-2026 | 502 (31%) | 474 (32%) | 498 (23%) | 1,127 (72%) |
| Birth Weight | 1,000 (750, 1,260) | 940 (720, 1,210) | 1,050 (750, 1,300) | 927 (660, 1,260) |
| Gestational Age | 28.00 (25.71, 30.14) | 27.29 (25.29, 29.43) | 28.71 (26.00, 31.00) | 27.57 (25.43, 29.57) |
| Died | 171 (11%) | 274 (18%) | 179 (8.3%) | 180 (11%) |
| Intraventricular Hemorrhage | 425 (26%) | 459 (31%) | 274 (13%) | 376 (24%) |
| Late Onset Sepsis | 258 (16%) | 187 (13%) | 143 (6.6%) | 165 (11%) |
| Necrotizing Enterocolitis | 16 (1.0%) | 13 (0.9%) | 41 (1.9%) | 19 (1.2%) |
| Intubated (qualified) | 335 (21%) | 146 (9.8%) | 123 (5.7%) | 772 (49%) |
| Any Vital Signs | 1,131 (70%) | 981 (66%) | 1,069 (49%) | 648 (41%) |
| Vital Signs (hours of life) | 1,291 (703, 2,122) | 212 (95, 565) | 852 (352, 1,611) | 1,470 (728, 2,195) |
| Vitals Coverage (%) | 96 (78, 99) | 15 (5, 57) | 73 (37, 96) | 94 (68, 99) |
| <sup>†</sup> n (%); Median (Q1, Q3) |  |  |  |  |

**Figure 1:**
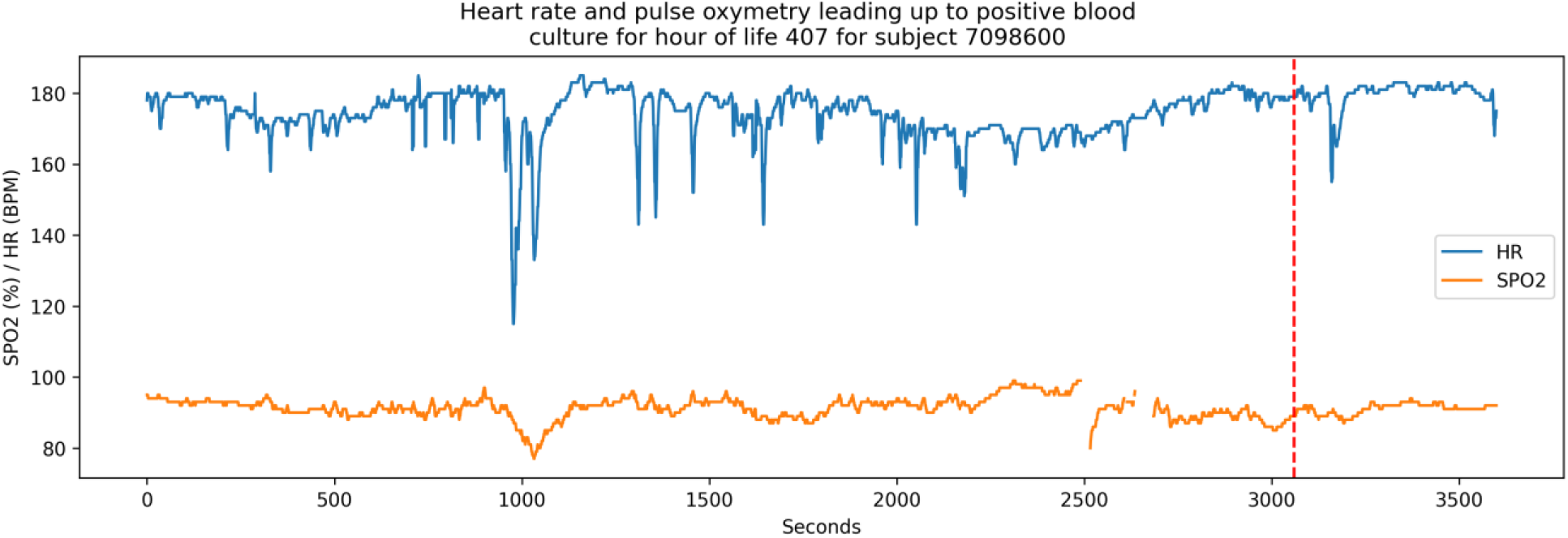
Plot of HR and SpO_2_ leading up to a positive sepsis culture (vertical red dashed line) for patient with *subject_number_deid* = 7098600. This plot was generated by the neurokit2 python package ^32^. A script to reproduce this figure is included in the dataset.

### Technical Validation

All data underwent systematic quality control procedures. Form completion status was tracked as incomplete (fields remaining to be completed), unverified (all available data entered but not reviewed), or complete (reviewed and confirmed). Site principal investigators audited at least 10% of subject entries and reviewed major outliers to ensure data accuracy. An ad-hoc visual comparison of 10-minute mean HR and SpO2 vital signs from randomly selected patients from this dataset showed nearly identical patterns as those in previously published data^33^.

### Usage Notes

Data are made available publicly as downloadable parquet files on the University of Virginia’s LibraData website^28^, itself a Dataverse instance (King, 2007). Within the Dataverse project, a scripts directory contains sample R, python, and MATLAB files for loading and merging REDCap and Vitals parquet files to produce images similar to the *gtsummary* table and segment plot contained herein.

## Data Availability

The PreMo dataset is available publicly on the University of Virginia’s LibraData website as a collection of parquet files, RO-Crate metadata, and sample script files^28^.

## Code availability

The deidentification and processing code is omitted to help prevent reidentification of protected health information. To illustrate reading and processing the dataset, sample scripts for plotting HR and SpO_2_ segments, creating a summary table, and producing the RO-Crate metadata are included within the dataset’s *scripts* directory.

## Notes

### Competing Interest Statement

Brynne A. Sullivan has received research support from Medical Prediction Sciences Corporation and Nihon Koden Digital Health Solutions. Doug E. Lake owns stock in Medical Prediction Sciences Corporation. Zachary A. Vesoulis has received research support from Medtronic, Edwards LifeSciences, and ReAlta Life Sciences, and has consulted for Medtronic. N. Ambalavanan is on the Data Safety Monitoring Board for Oak Hill Bio LLC and is a medical advisor for ResBiotic and AlveolusBio. Colm Travers is supported by a grant from the National Heart Lung and Blood Institute (K23 HL157618), and is funded by research grants from Owlet Baby Care Inc. and Prapela Inc. No authors have any other financial or non-financial conflicts of interest.

### Funding Statement

We acknowledge the following grant for funding the work presented in this manuscript: R01 HD072071 [Co-PIs KD Fairchild & BS Sullivan].

### Author Declarations

IRB of Columbia University gave ethical approval for this work in #IRB-AAAP5807 for retrospective analysis with waiver of consent IRB of University of Alabama at Birmingham gave ethical approval for this work in #IRB-300007524 for retrospective analysis with waiver of consent IRB of University of Virginia gave ethical approval for this work in IRB-HSR #21237 for retrospective analysis with waiver of consent IRB of The Washington University in St. Louis gave ethical approval for this work in #201803222 for retrospective analysis with waiver of consent

